# Trend prediction of COVID-19 based on ARIMA model in mainland of China

**DOI:** 10.1101/2020.09.04.20188235

**Authors:** Chuqiao Han, Xifeng Ju, Jianghua Zheng

## Abstract

The ongoing pandemic of COVID-19 has aroused widespread concern around the world and poses a severe threat to public health worldwide. In this paper, the autoregressive integrated moving average (ARIMA) model was used to predict the epidemic trend of COVID-19 in mainland of China. We collected the cumulative cases, cumulative deaths, and cumulative recovery in mainland of China from January 20 to June 30, 2020, and divided the data into experimental group and test group. The ARIMA model was fitted with the experimental group data, and the optimal model was selected for prediction analysis. The predicted data were compared with the test group. The average relative errors of actual cumulative cases, deaths, recovery and predicted values in each province are between −22.32%−22.66%, −9.52%−0.08%, −8.84%−1.16, the results of the comprehensive experimental group and test group show The error of fitting and prediction is small, the degree of fitting is good, the model supports and is suitable for the prediction of the epidemic situation, which has practical guiding significance for the prevention and control of the epidemic situation.

**Highlights:**

1. We predicted future COVID-19 occurrences in mainland of China based on ARIMA model.
2. We validated the model based on the previous outbreak data with actual data for June, 2020.
3. The measures taken by the government have contained spread of the epidemic
4. The combination of multiple models may improve the robustness of the model

## 1. Introduction

COVID-19 has spread worldwide and has caused tens of thousands of human deaths. At the same time[1], it has a huge impact on the world’s medical, public health, economic, and other aspects [2]. In order to effectively suppress the spread of COVID-19, the Chinese government has successively announced the launch of a first-level response mechanism for major public health emergencies and implemented strict prevention and control measures[3]. By the end of June 2020, mainland of China had 85000 confirmed COVID-19 cases and 4648 deaths. Although the situation of prevention and control is getting better, the global situation is still not optimistic.

ARIMA model considers the law that the historical data of the research object itself changes with time, and uses this to predict future values, that is, to replace various influence factors with time. At present, the advantage of ARIMA model in predicting infectious diseases has been confirmed in many studies[4–5].

This study is based on the autoregressive integrated moving average model to predict the cumulative cases, cumulative deaths, and cumulative recovery of COVID-19 in mainland of China, and select the most appropriate model to simulate the epidemic pattern by simulating multiple models. A preliminary explanation is given to evaluate the effects of the epidemic prevention and control measures at this stage.

## 2. Data and Methods

### 2.1 Data Sources

The COVID-19 case data comes from the National and local health and construction commission daily information release (https://ncov.dxy.cn/). We collected the cumulative cases, deaths, recovery in mainland of China from January 20, 2020 to June 30, 2020.

### 2.2 Methods

ARMA model is a stationary time series model, but COVID-19 daily cases, deaths and recovery changes with volatility, uncertainty, is a non-stationary random process, so this article introduces the difference operator I (The original non-stationary time series can be improved into a stationary time series after d-order difference), establish the COVID-19 cumulative cases, deaths, and recovery trend prediction ARIMA model. In recent years, the ARIMA model has become one of the most commonly used methods in the prediction of many epidemics. This method is particularly suitable for short-term prediction of infectious diseases, and its prediction accuracy has been widely recognized, at the same time, it can also provide effective help for disease prevention and policy-making[6].

ARIMA model is composed of autoregressive model (AR model (P)), moving average model(MA model (q)) and difference operator I (d), where p, d and q are autoregressive order, difference order and moving average order of time series[7]. The expression of the p-order AR model is as follows:

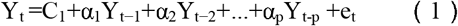

Where αi (i = 1,2,…,p) is the AR model coefficient, e_t_ is the random interference term, and C_1_ is the constant.

The mathematical expression of the q order MA model is:

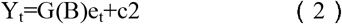

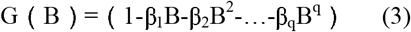

Where β_j_ (j = 1,2,…,q) is the MA model coefficient, e_t_ is the random interference term, and c_2_ is the constant.

Combining the time series after the difference of order d, the final expression of the ARIMA model is:

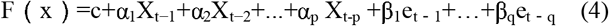

Where α represents the coefficient of AR, β represents the coefficient of MA, and c is the constant term.

ARMA model construction (Fig. 1 The flowchart of ARIMA model|Fig.1):

**Fig.1.**
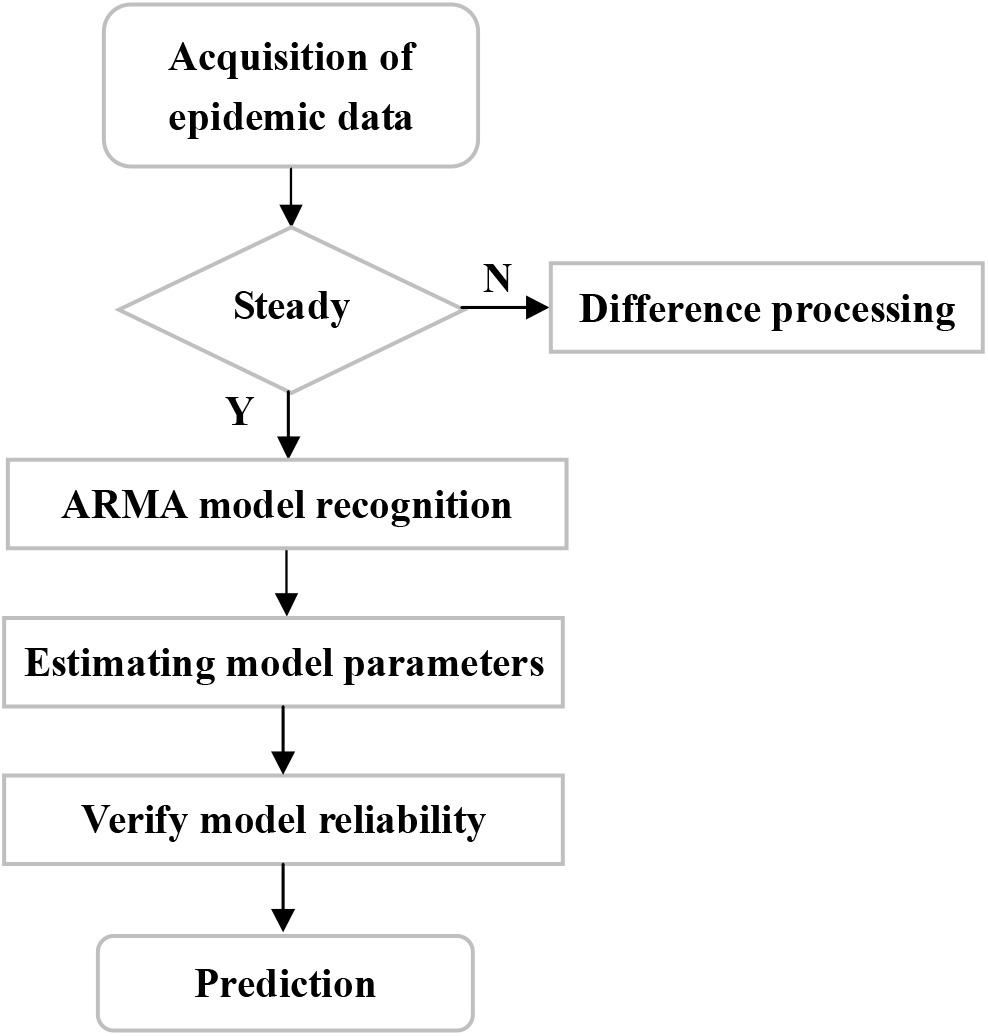
The flowchart of ARIMA model

(1) Data difference: the collected COVID-19 data are divided for several times to make it stable, and then the autocorrelation and partial autocorrelation tests are performed on the differential data.

(2) The choice of modeling parameters: according to an information criterion (AIC) and Bayesian information criterions (BIC), combing with the autocorrelation function (ACF) and partial autocorrelation function (PACF) of the residual sequence to determine the order of the model, and finally select the model with the highest fitting degree[8].

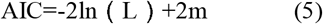

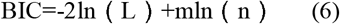

Where: L is the maximum likelihood function of the model, m is the number of estimated parameters, and n is the sample size.

(3) Verify the reliability of the model: we use the model to predict the cumulative cases, deaths, recovery in Chinese provinces (excluding Hong Kong, Macao, and Taiwan) from June 1-30, 2020, calculate the relative error from the actual cases, and verify the model predictive effect.

## 3. Results

We build a model based on the cumulative cases, deaths, and recovery data in mainland of China from January 20 to May 30, 2020. Here we take Hubei Province as an example to describe the recognition process of the model.

First, we build the original time series figure (Fig.2), from the figure, we can see that the initial outbreak of COVID-19 increased exponentially. The Chinese government took timely measures to control the epidemic. It can be seen from Fig. 2 that the original data time series are not stationary, so we need to carry out first-order differential processing on the original data. The processed time series is shown in Fig. 3. Since the time series in Fig. 3 is still unstable, we need to make two difference, d = 2, as shown in Fig. 4. It can be seen from the figure that the data floats up and down in “0”, which can judge that the data is stable after the second-order difference, so d = 2 is determined.

**Fig.2.**
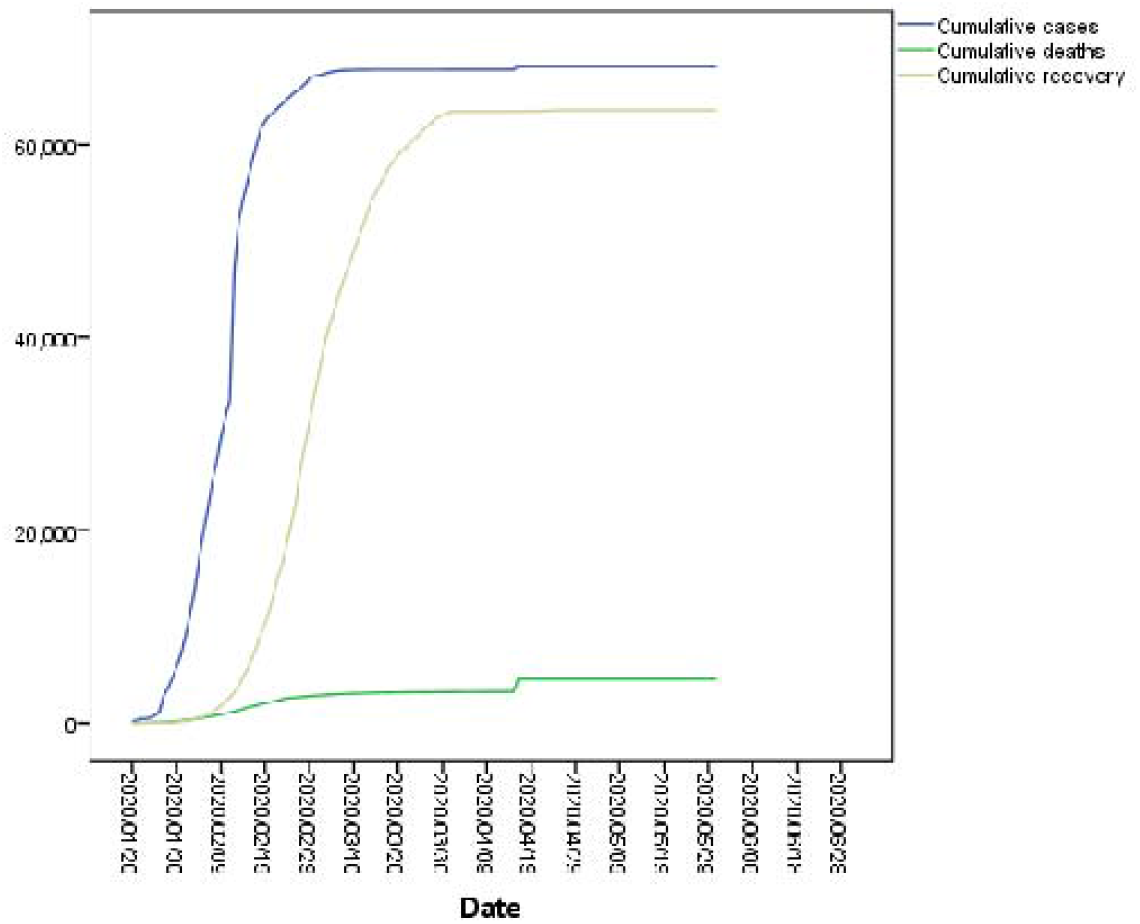
The cumulative cases, deaths, recovery original time series figure

**Fig.3.**
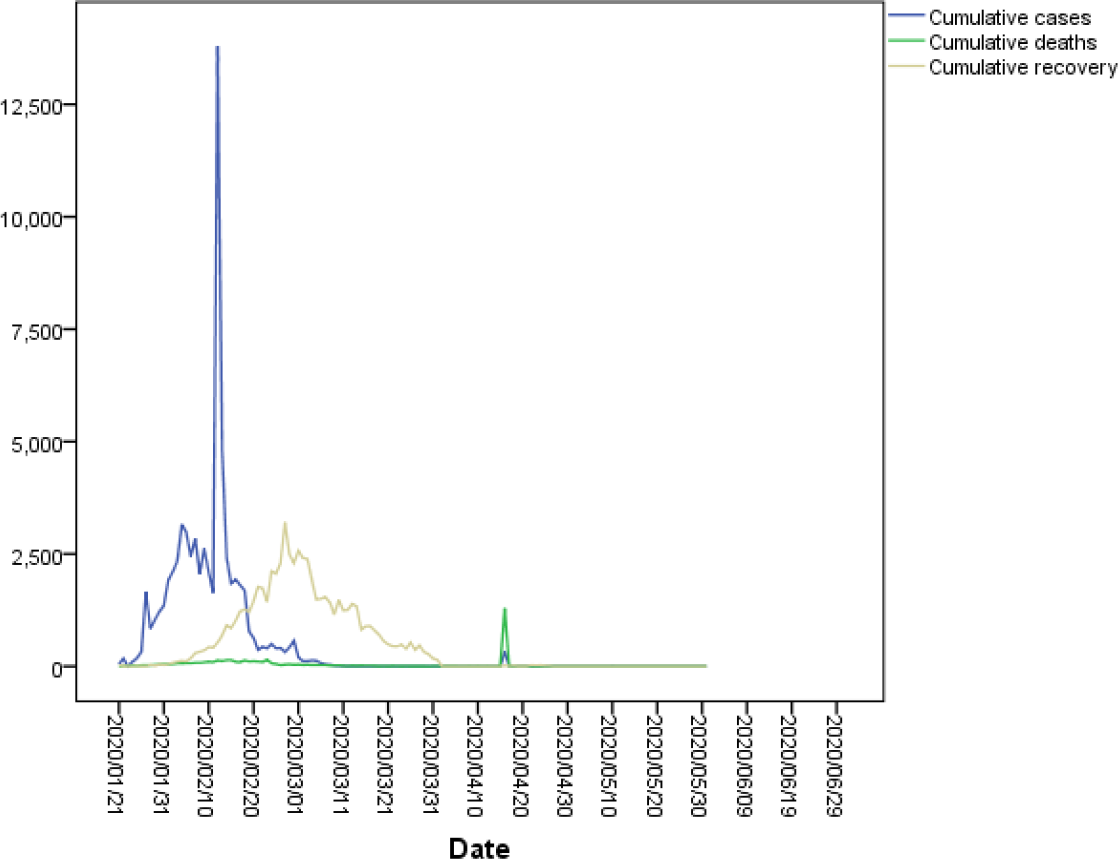
First-order difference processing time series figure (d = 1)

**Fig.4.**
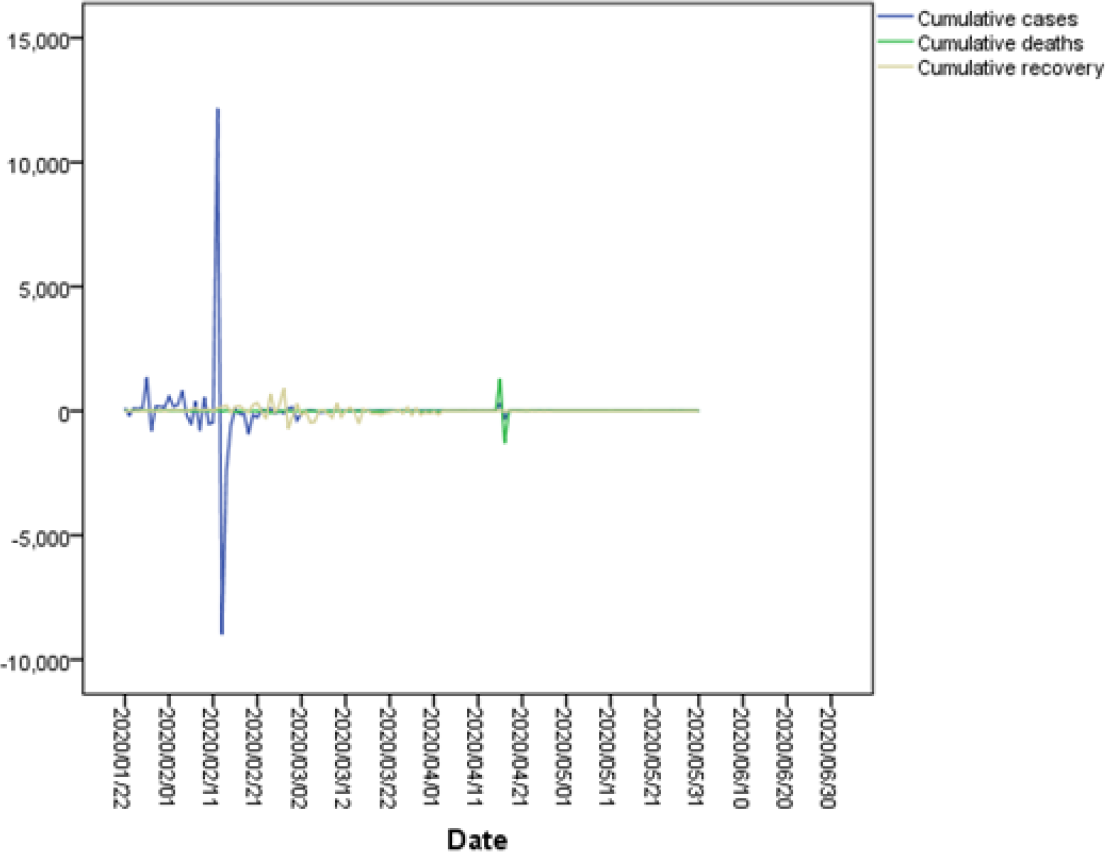
Second-order difference processing time series figure (d = 2)

After the d value is determined to be 2 by differential processing, we need to select different combinations of p and q values to test the goodness of fit, and use the models with relatively small AIC and BIC values as the best model, and then verify the model.

In the end, we obtained the models for predicting the cumulative cases, deaths, and recovery in Hubei Province as ARIMA(0,2,1), ARIMA(1,2,1), ARIMA(1,2,4). The residual of the model is tested by white noise sequence. As shown in Fig. 5, the autocorrelation function and partial autocorrelation function of the residual sequence are basically within 95% confidence interval, indicating that there is no autocorrelation in the residual sequence. Then the model through the white noise test, the model can be used to predict the cumulative cases, deaths, and recovery in Hubei Province.

**Fig.5.**
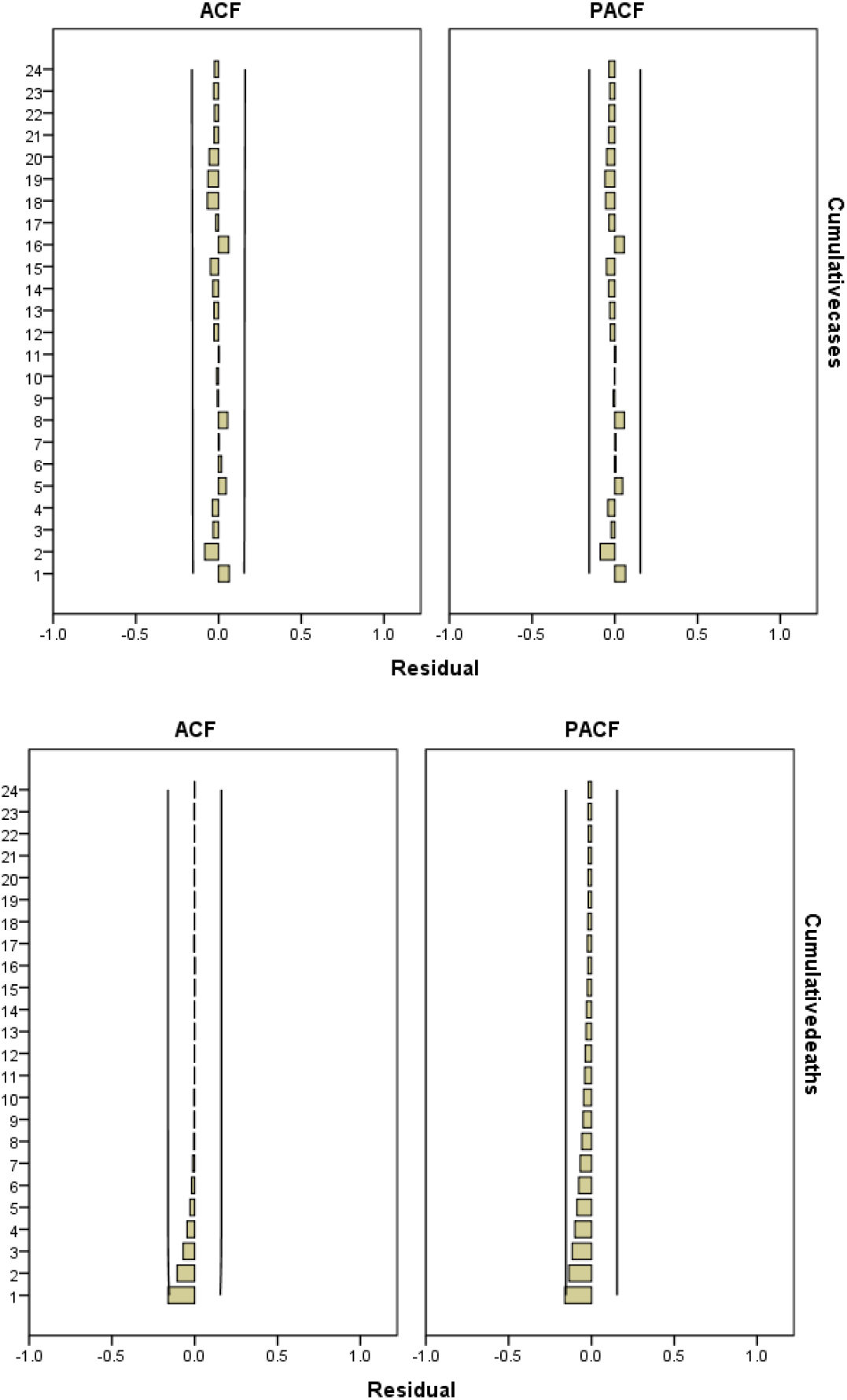
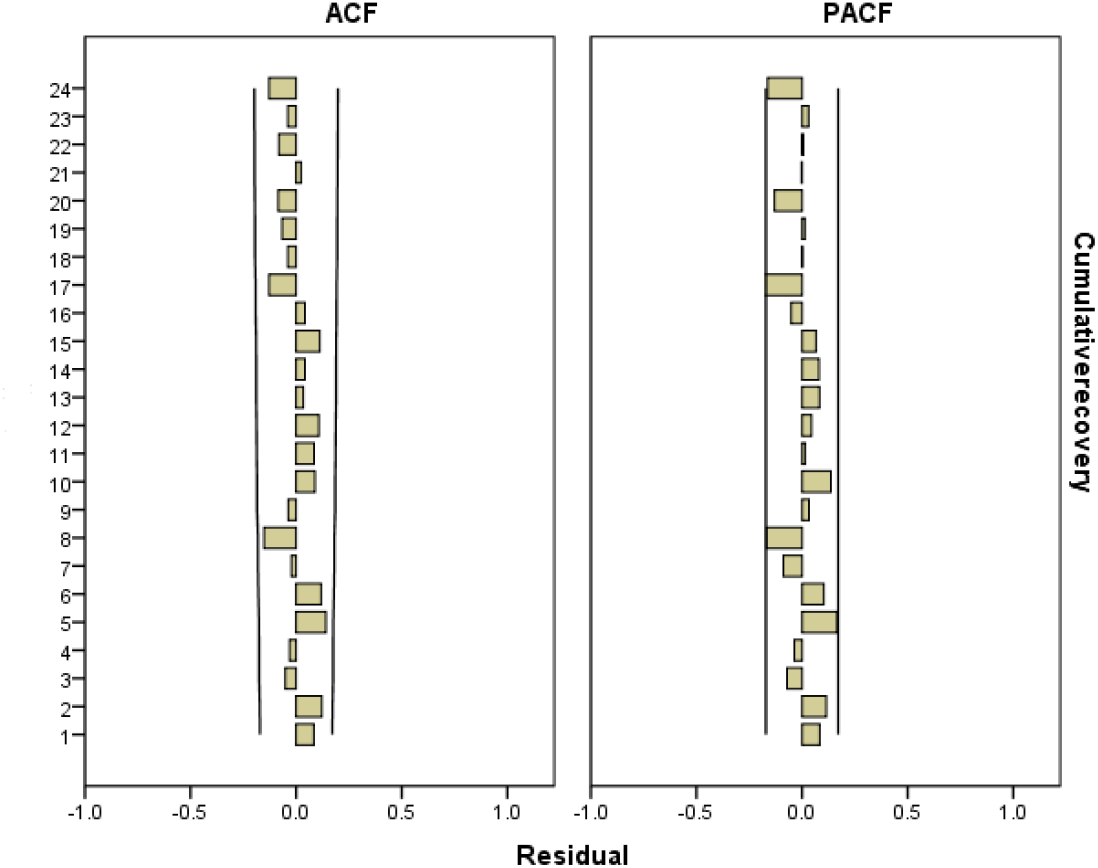
Residual sequence figure of ARIMA model for the cumulative cases, deaths, recovery in Hubei Province

Model test and error analysis: We divided the data into experimental group (January 20 - May 31, 2020) and test group (June 1 - June 30, 2020), First of all, we bring the data of the experimental group into the ARIMA model of the cumulative cases, deaths, recovery in each province, and calculate the average relative error between the actual number and the fitted value. The results are shown in Table 1.

**Table 1.**
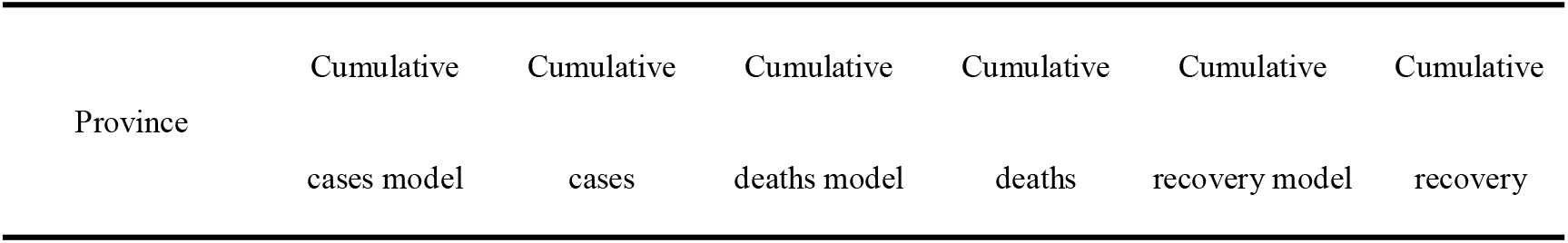
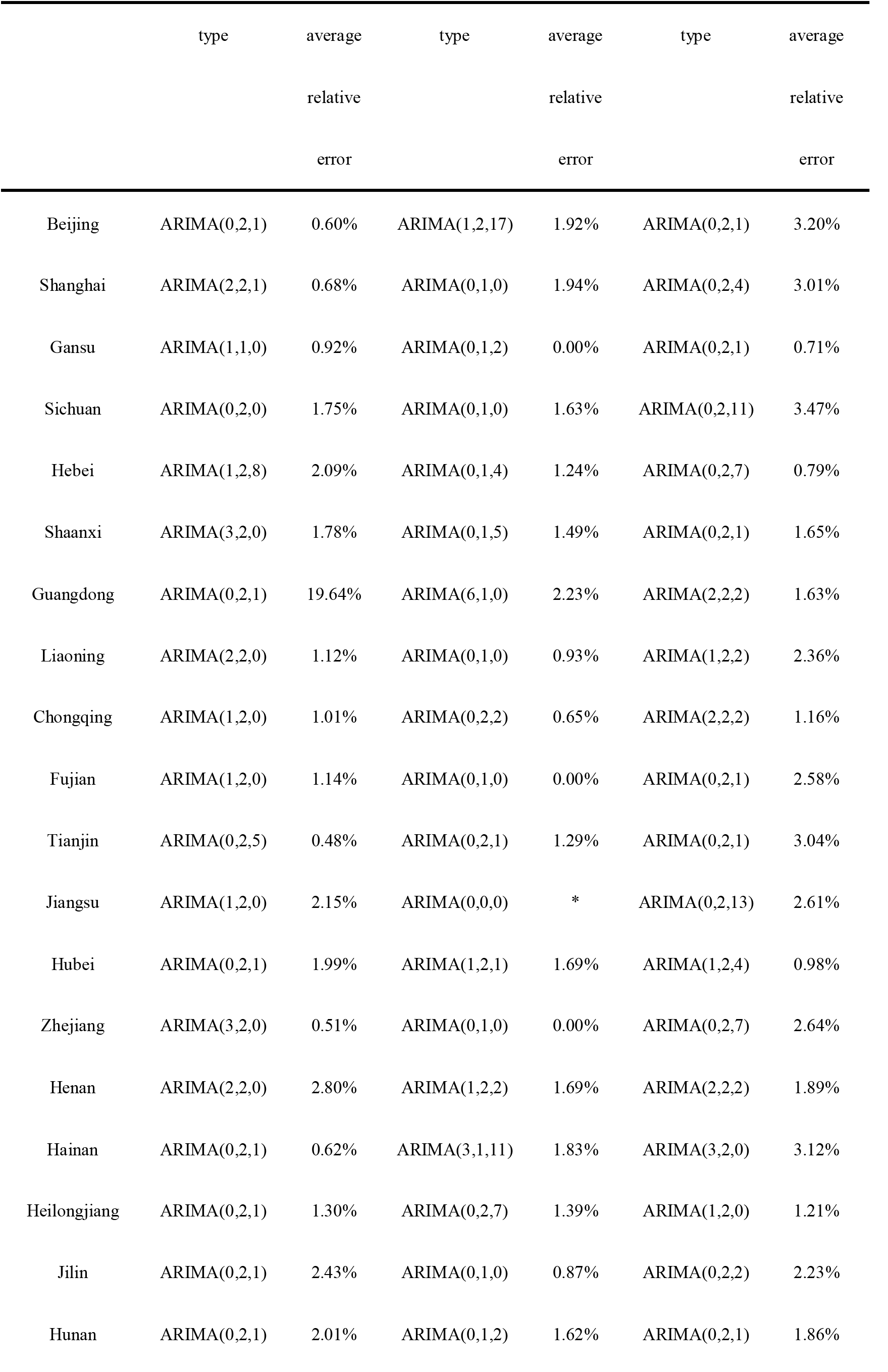
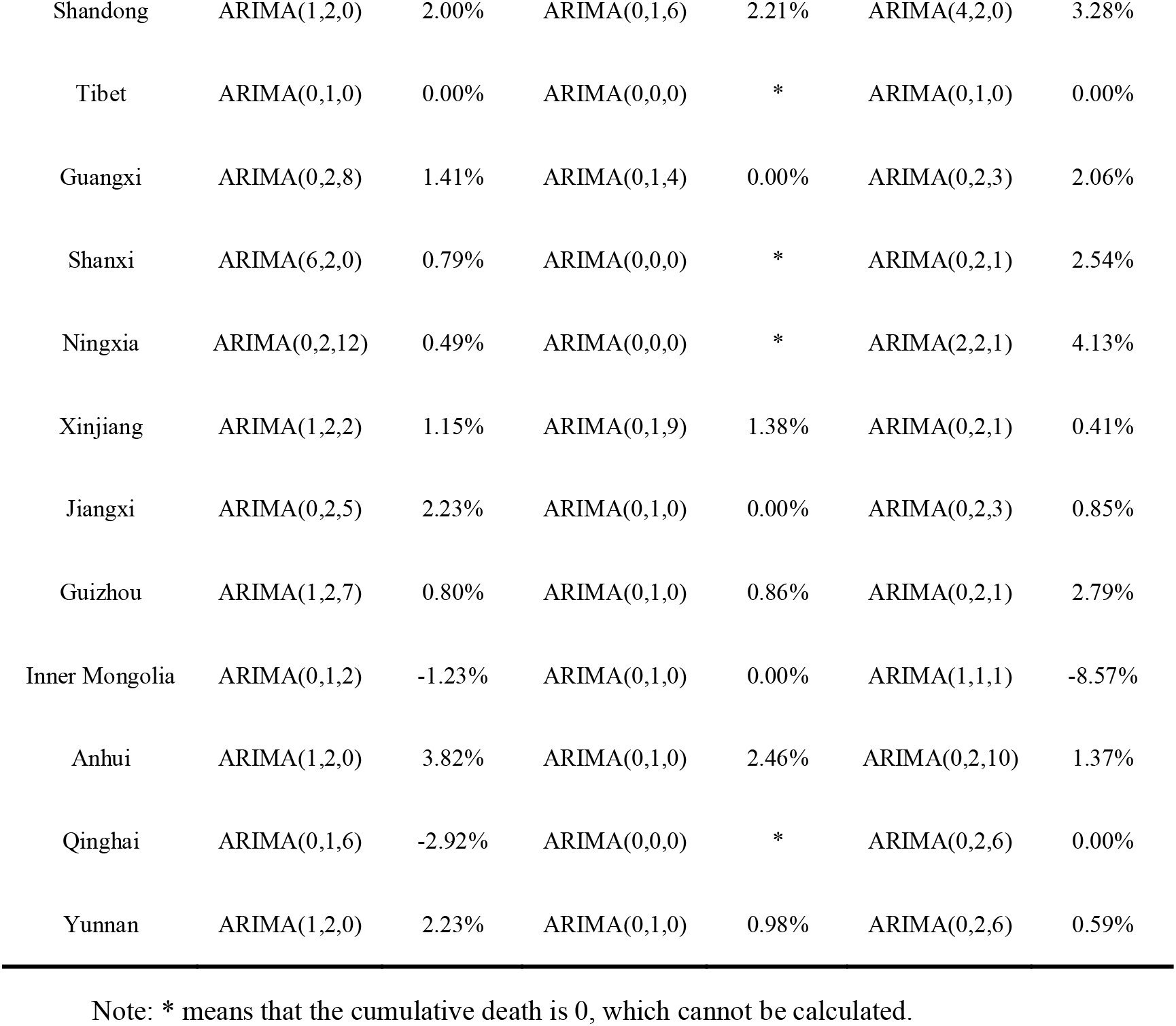
the average relative error table of actual cumulative cases, deaths, recovery and fitted values in each province during January 20, 2020 to May 31, 2020

It can be seen from Table 1 that the average relative error of the cumulative cases in each province is between −2.92% and 3.82% (except for Guangdong Province); the average relative error of the cumulative cases in Guangdong Province is 19.64%. By analyzing the data of Guangdong Province, we found that on January 23, 2020, the actual cumulative cases were 26, and the fitting value was 1. This is mainly since at the early stage of the epidemic, the transmission route of COVID-19 was not known, and the government did not take effective measures, so the number of cases increased exponentially, which led to a large error at that time. The average relative errors of cumulative deaths and recovery in various provinces are between 0-2.46% and −8.57%-4.13%, the errors are small and the fit is good, so the model is suitable. We further analyzed the average relative error between the actual cases and the predicted values in each province of the test group (June 1 - June 30, 2020). The results are shown in Table 2.

**Table 2.** the average relative error table of actual cumulative cases, deaths, recovery and fitted values in each province from June 1st to 30th

| Province | Cumulative cases | Cumulative deaths | Cumulative recovery |
| --- | --- | --- | --- |
|  | average relative error | average relative error | average relative error |
| Beijing | 22.66% | 0.00% | 0.45% |
| Shanghai | 3.00% | -9.40% | -0.50% |
| Gansu | 6.47% | 0.00% | 0.66% |
| Sichuan | -21.32% | -6.67% | -0.34% |
| Hebei | 2.76% | 0.00% | 0.60% |
| Shaanxi | -0.32% | 0.00% | 0.19% |
| Guangdong | 0.44% | 0.00% | 1.16% |
| Liaoning | 1.74% | 0.00% | -0.98% |
| Chongqing | 0.30% | 0.00% | 0.00% |
| Fujian | 0.88% | 0.00% | -2.07% |
| Tianjin | 1.93% | 0.00% | 1.10% |
| Jiangsu | 0.05% | * | 0.26% |
| Hubei | 0.56% | 0.08% | 0.05% |
| Zhejiang | 0.04% | 0.00% | 0.00% |
| Henan | 0.00% | 0.00% | -0.28% |
| Hainan | 0.87% | 0.00% | 0.60% |
| Heilongjiang | 0.21% | -8.70% | 0.21% |
| Jilin | -0.15% | 0.00% | -8.84% |
| Hunan | 0.00% | 0.00% | 0.00% |
| Shandong | -1.30% | 0.00% | 0.35% |
| Tibet | 0.00% | * | 0.00% |
| Guangxi | -0.21% | 0.00% | 0.00% |
| Shanxi | 0.00% | * | 0.00% |
| Ningxia | 0.00% | * | 0.00% |
| Xinjiang | 0.00% | 0.00% | 0.00% |
| Jiangxi | 0.00% | 0.00% | 0.00% |
| Guizhou | 0.00% | 0.00% | 0.00% |
| Inner Mongolia | -9.13% | 0.00% | 0.62% |
| Anhui | 0.00% | -9.52% | 0.00% |
| Qinghai | 0.00% | * | 0.00% |
| Yunnan | 0.00% | 0.00% | 0.44% |

Note: \* means that the cumulative death is 0, which cannot be predicted.

It can be seen from Table 2 that the average relative errors of actual cumulative cases, deaths, recovery and predicted values in each province are between −22.32%-22.66%, −9.52%-0.08%, −8.84%-1.16%. The results of comprehensive experimental group and test group show that the error between fitting and prediction is small, the fitting effect is good, the model supports and is suitable for epidemic situation prediction.

COVID-19 trend in mainland of China: According to each provincial model, we get the overall trend of COVID-19 in mainland of China, as shown in Fig. 6. The trend of the fitting line between the predicted value and the actual value of the cumulative cases, deaths and recovery was consistent (6a, 6b, 6c). At the beginning of the epidemic, the cases showed an exponential upward trend. At the end of January, the Chinese government took measures to seal the city, isolate it from the source of infection, and reduce the possibility of infection, the epidemic was controlled in mainland of China. The epidemic situation in Hubei, Guangdong, Zhejiang, Henan and other places in mainland of China is relatively serious (6a). Residents in this area still need to do epidemic prevention and control work to prevent the possibility of a second rebound of the epidemic situation.

**Fig.6.**
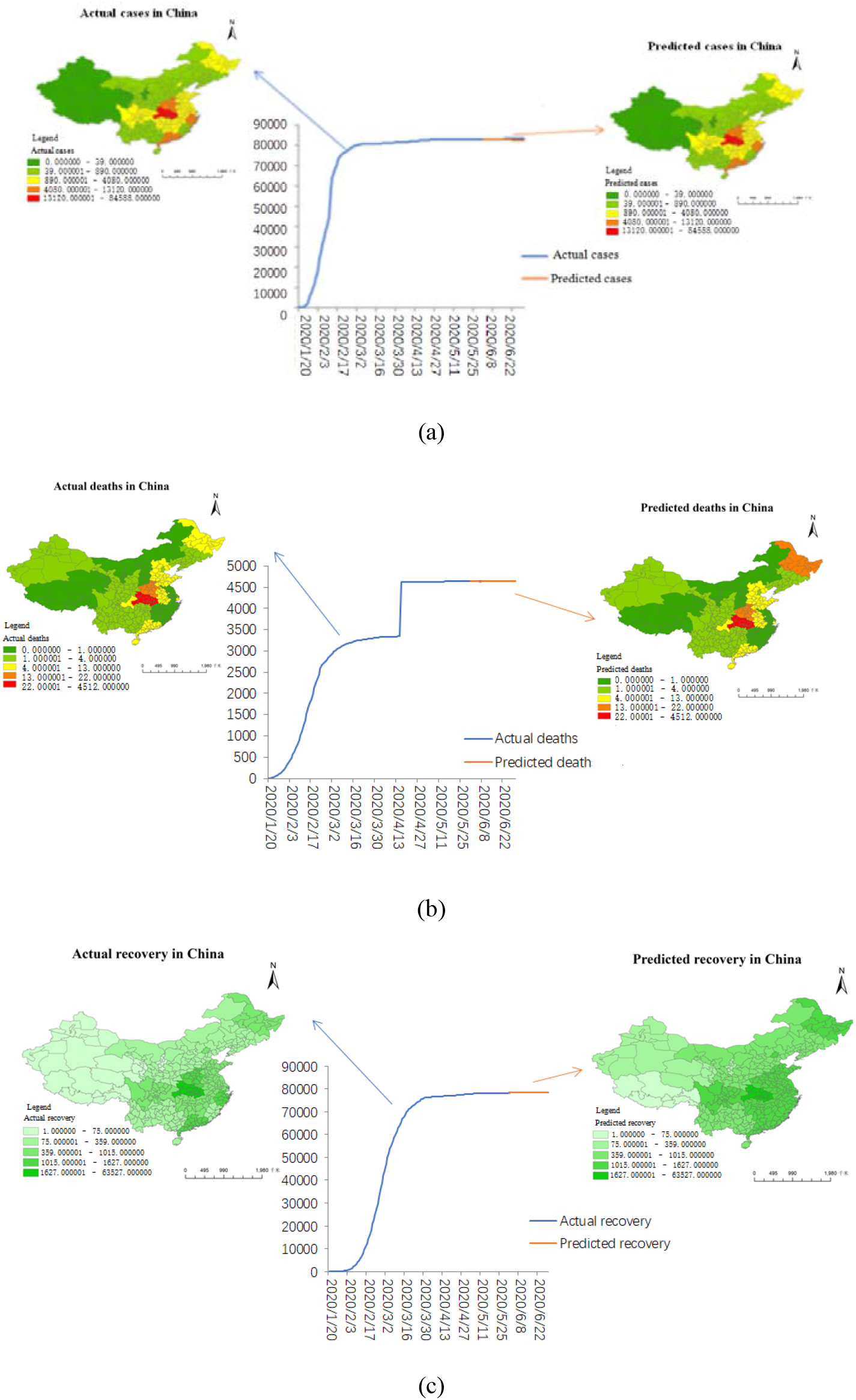
Cumulative cases, deaths, recovery figure in mainland of China

With the increase of confirmed cases, the number of deaths initially gradually increased. It can be seen from 6b that the number of deaths in mid-April showed a linear upward trend. The reason for this data surge is that the preliminary data statistics are incomplete, the statistical standards are inconsistent and there are varying degrees of delay in the statistical process [9]. The state has proposed efforts to reduce deaths, requiring provinces to increase the number of ventilators to ensure adequate medical facilities. The number of deaths in mainland of China has remained stable around May. In this fight against the epidemic, many volunteers have appeared in many places, and medical personnel throughout the country have conducted targeted research on the epidemic, and their experience and level have been improved, resulting in the continuous improvement of the number of cures (6c).

## 4.Discussion

Data from January to June 2020 shows that the epidemic in mainland of China has peaked at the end of February and has been steadily declining since then. This is mainly due to the closed management measures taken by the Chinese government in the face of the COVID-19 pandemic to stop population-intensive Place, and achieved good results. By the end of March 2020, all regions had lifted the closure measures and adjusted the primary response to major public health emergencies to secondary, and the cases in the epidemic have not changed significantly since then. This may be related to climate factors such as continuous temperature increase in various places. The studies have shown that changes in temperature and humidity may be important factors affecting the spread of COVID-19[10].

In recent years, ARIMA model, neural network and so on are more active in the field of epidemic prediction because they can relatively accurately explore the occurrence and development of epidemics[11–15].

The reason why ARIMA model can be widely favored in the field of epidemiological prediction is that it not only absorbs the advantages of traditional regression analysis but also takes advantage of the moving average, and can affect many factors affecting the development of the epidemic (such as temperature, humidity, aerosol, population migration, etc.) integrated into time variables for quantitative expression, is a method with strong practicability and high prediction accuracy. However, ARIMA model also has certain limitations. It is a mathematical model built on past historical data. Therefore, ARIMA model is only suitable for short-term forecasting. If the forecasting time is too long, it will increase the forecasting error and affect the forecasting accuracy[16–18].

At present, COVID-19 has become a global epidemic, and the epidemic situation is still accelerating and has not yet reached its peak. The prevention and control of the epidemic situation must not be delayed. Based on the COVID-19 data of mainland of China, we found that the model has a high degree of fitting, which can predict the development trend of the epidemic situation in the future. However, in practice, other factors[19–22] will also affect the development trend of the epidemic situation, causing predictions to deviate, this needs further study. In practical applications, we can make the model have better prediction effect and accuracy by combining with multiple models.

## 5.Conclusion

We used the epidemic data of Chinese provinces from January 20 to June 31, 2020, in which the data before May 30 were involved in model fitting, the data of June were tested in the model, and the ARIMA model was used to fit the data, so as to obtain the epidemic prediction of different periods in mainland of China. Through the verification of the existing data, the prediction effect is good. The model can be used to predict the epidemic situation in mainland of China in the future and make positive contributions to policy makers’ prevention and control of the epidemic and protection of people’s lives. On the other hand, the applicability and robustness of the model in mainland of China can also be studied in other countries and regions, so as to verify the accuracy and improve the performance of the model and provide assistance for epidemic prevention and control in this region.

## Data Availability

The COVID-19 case data comes from the National and local health and construction commission daily information release (https://ncov.dxy.cn/).

## Acknowledgements

This work was financially supported by spatial data analysis and geoscientific calculation are used to study urban vulnerability based on microgeographic units(grant numbers: 41461035). Thanks to the National and local health and construction commission for providing data on the daily cumulative cases, deaths and recovery.

